# CalPred yields calibrated intervals for polygenic risk prediction

**DOI:** 10.64898/2026.04.21.26351410

**Authors:** Zhuozheng Shi, Zixuan (Eleanor) Zhang, Ravi Mandla, Kangcheng Hou, Bogdan Pasaniuc

## Abstract

Polygenic scores (PGS) have emerged as a useful biomarker for stratification of high-risk individuals in genomic medicine, with prediction intervals arising as a principled approach to incorporate statistical uncertainty in their individual-level predictions. In contrast to recent reports by Xu et al^7^, we show that CalPred^6^ provides well-calibrated prediction intervals that contain the trait phenotypes at targeted confidence levels. CalPred maintains calibration when PGS performance varies across contextual factors (e.g., ancestry, age, sex, or socio-economic factors) whereas PredInterval^7^ – a recently introduced method that focuses on marginal calibration across all individuals – exhibits miscalibration.

## Main

Polygenic scores (PGS) have emerged as a principled approach for stratification of at-risk patients in precision medicine^1,2^. However, PGS-based predictions carry high levels of statistical noise at the individual level^3–5^, underscoring the need for confidence intervals to accurately reflect the range of an individual’s predicted genetic risk profile. Indeed, two recently introduced approaches (CalPred^6^ and PredInterval^7^) provide individual-level prediction intervals for PGS-based prediction, with Xu et al.^7^ claiming PredInterval is the only approach that maintains calibration in simulation and real data. We show that Xu et al.’s CalPred conclusions arise from not separating data used for training versus calibration thus violating a basic principle of model evaluation which renders the CalPred results of Xu et al uninterpretable. We confirm with Xu et al authors that correct use of CalPred, ensuring independence between training and testing (usually done by reserving a fraction of training data for calibration), leads to calibration across all simulation scenarios. Next, motivated by the pervasive context-specificity in PGS performance^5,8–12^ (e.g., PGS performance varies with ancestry, age, sex for most existing PGSes), we investigate contextual calibration of existing methods. We find that CalPred is the only approach that maintains calibration across contexts such as age, sex, or socio-economic status.

### CalPred prediction intervals are calibrated in simulations

We replicate the simulation scenarios of Xu et al to show that CalPred is calibrated at all settings, in contrast to results reported by Xu et al. CalPred achieved the targeted 95% coverage across all scenarios with varying heritability (20%-80%) and polygenicity (0.1-100% causal variants; **Figure 1a**) and maintained calibration across confidence levels from 20% to 99% **(Figure 1b)**. CalPred has comparable high-risk identification rates to PredInterval while achieving lower false-positive rates **(Figure 1c,d)**. PredInterval yielded wider intervals than CalPred **(Supplementary Figure 1)**, leading to higher false-positive rates (proportion of low-risk individuals incorrectly classified as high-risk) across thresholds defining low-risk individuals (bottom 5%, 25%, and 50%). As a result, PredInterval misclassified 1.39-1.42 times more low-risk individuals as high-risk compared to CalPred **(Figure 1e)**.

**Figure 1.**
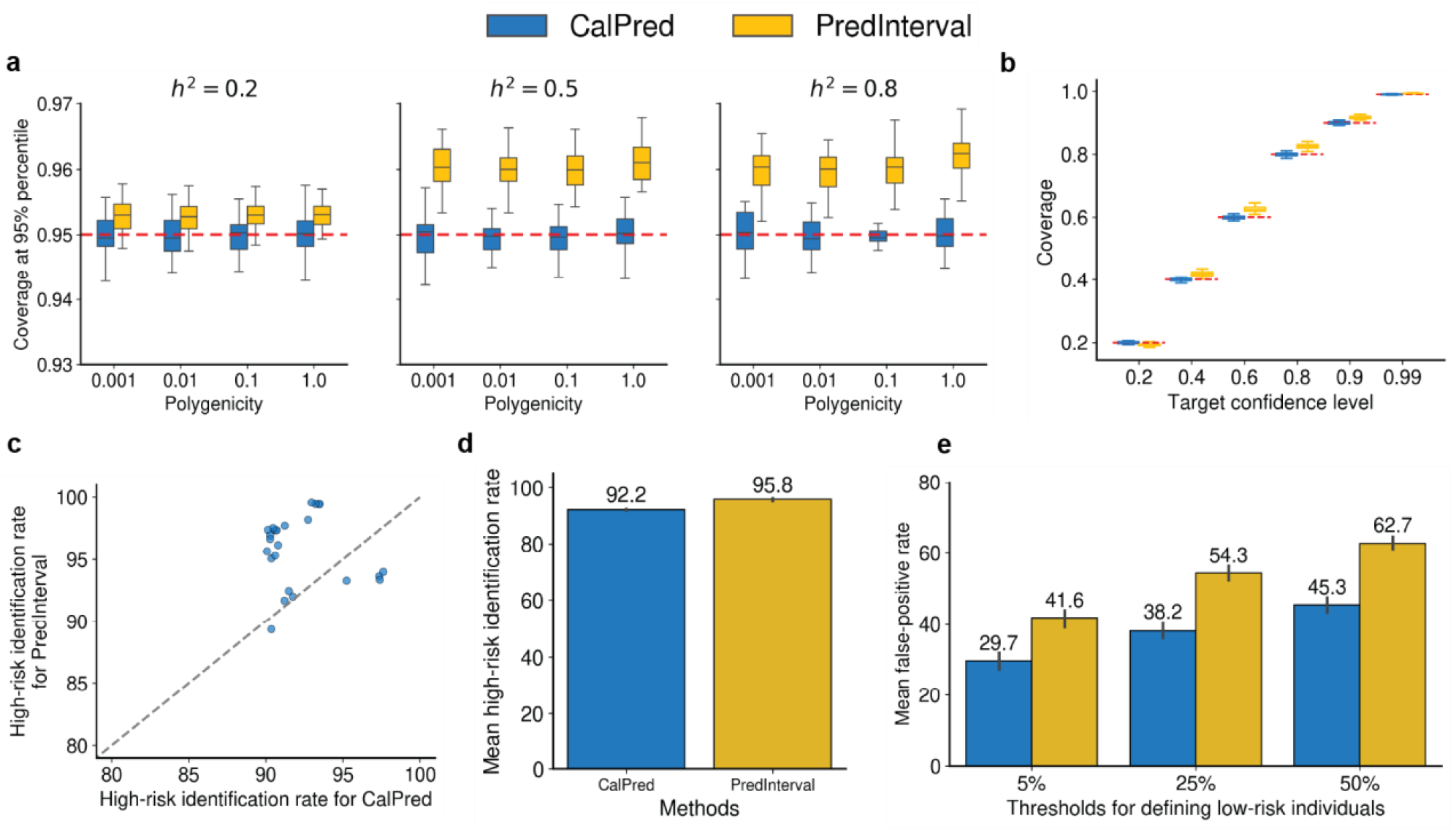
CalPred is calibrated across simulated scenarios by Xu et al. We simulated quantitative traits across varying polygenicity and compared the performance of CalPred (blue) and PredInterval (yellow). Each simulation setting was repeated 30 times. **a-b**. Results using a training dataset of size 50,000. The red dashed line indicates the targeted confidence level. **a**. Coverage at the 95% confidence level across simulation settings with varying polygenicity = 0.001, 0.01, 0.1, and 1, and heritability = 0.2, 0.5, and 0.8. **b**. Coverage across targeted confidence levels 0.2, 0.4, 0.6, 0.8, 0.9, and 0.99, with heritability of 0.5 and polygenicity of 1. **c-e**. Evaluation of high-risk classification performance across 24 simulation settings, including the 12 settings shown in a,b, and an additional 12 settings with identical heritability and polygenicity but with the training dataset size of 5,000. **c**. Comparison of high-risk identification rates between CalPred and PredInterval; each point represents a simulation setting. **d**. Mean high-risk identification rate across all 24 simulation settings. **e**. Mean false positive rate, shown across three thresholds, 5%, 25%, and 50%, used to define low-risk individuals.

Having shown that CalPred is calibrated, we next investigated the source of discrepancy in results reported by Xu et al. In Figure 1 above CalPred and PredInterval are provided with the same training and testing data, with CalPred reserving a subset of the training set as independent calibration data per its guidelines and as described in the CalPred manuscript^6^ (**Figure 1, Methods**). We identified the discrepancy in the CalPred results reported by Xu et al in that they used the same data for training and calibration for CalPred, thus breaking the independence assumption between the datasets and leading to overfitting. Indeed, when we provide the same calibration and training data to CalPred leading to overfitting, we observed similar miscalibration results (CalPred*-overfit; **Supplementary Figure 2, Methods**). This underscores the need for independence between training and calibration data for CalPred to achieve calibration with future work investigating the size of the calibration data as a function of considered contexts.

### CalPred is calibrated in the presence of context-specific PGS performance

Results above assume uniform PGS prediction performance across individuals – a simplification that rarely holds in practice; PGS accuracy varies substantially across contexts such as genetic ancestry, age, sex, and other socio-demographic features^5,8–13^. When PGS prediction accuracy varies with context, calibration averaged across all individuals (marginal calibration) provides no guarantee of calibration within a specific context – for example, the 95% prediction intervals could, on **average**, contain 95% of the true phenotype values across individuals across age strata, whereas they could be too narrow for a specific age strata (e.g., over 60) thus leading to increased false positive rate for that context when used in risk stratification. We explored the impact of PGS contextual accuracy on calibration in simulations (**Methods**) to find that CalPred provided 95% calibrated prediction intervals across all modeled context deciles and all PGS performance variability magnitudes. In contrast, PredInterval is increasingly mis-calibrated at extremes of context as PGS contextual prediction (ΔR^2^) increased (**Figure 2**). For example, at a ΔR^2^ of 150%, PredInterval’s miscalibration at the 50% confidence level increases from 5% to 25% in the bottom context decile and from 11% to 26% in the top. This is expected as PredInterval does not incorporate context into its approach, yielding static prediction intervals that are constant across all individuals (**Supplementary Figure 3**). For completeness, after consulting Xu et al authors in February 2025, we also evaluated a version of PredInterval where context and covariates are added in the model fitting to find similar results **(Methods; Supplementary Figure 4)**. This is expected as PredInterval with PGS+Covariates models context only through the mean and does not account for context-specificity in phenotypic variance. Together, these results demonstrate that marginal (average) calibration may under-represent calibration within specific population subgroups — a critical limitation for PGS implementation^1,2,5^.

**Figure 2.**
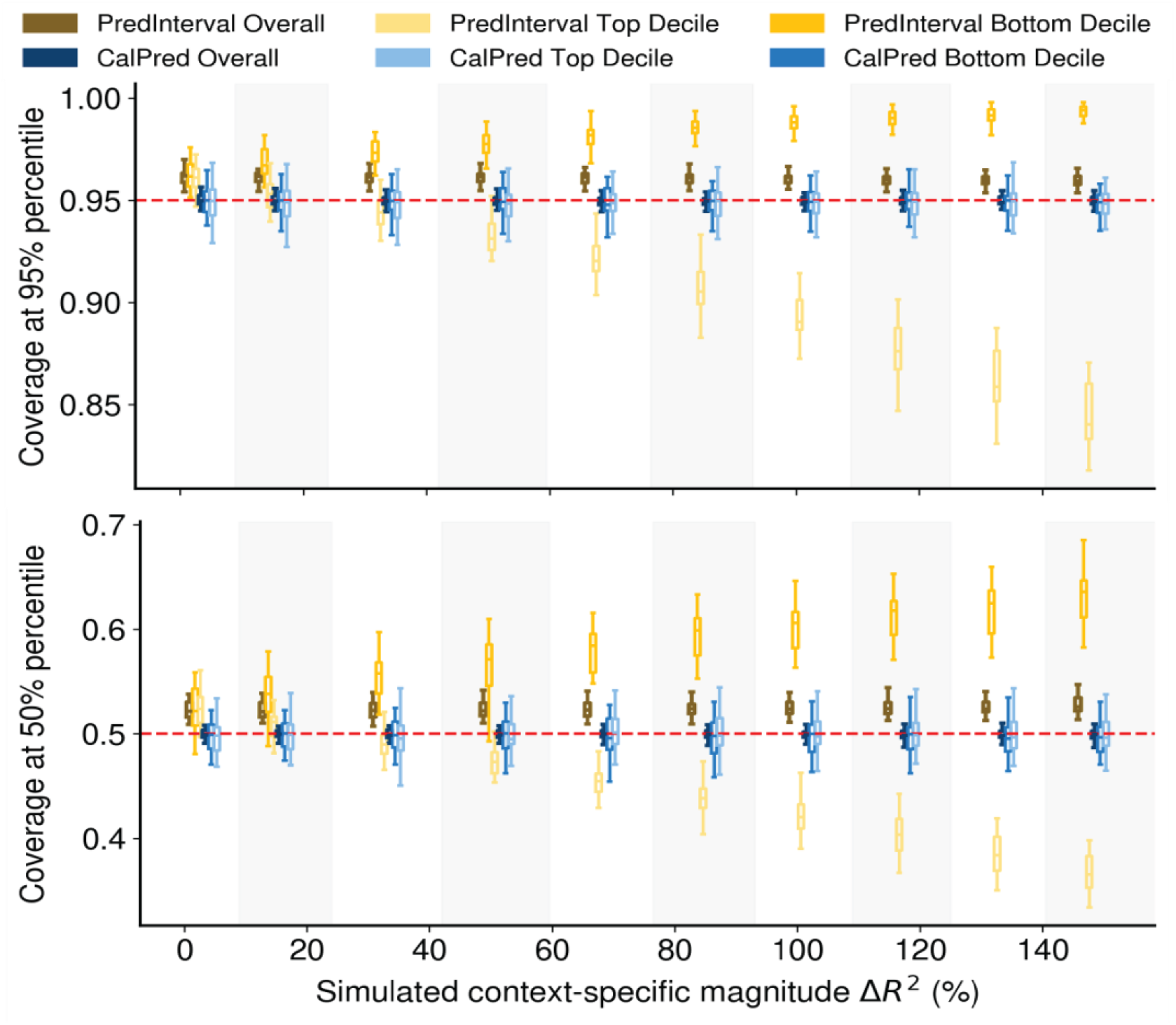
CalPred is calibrated in the presence of context-specific effects on PGS accuracy. We compared the coverages of PredInterval (yellow) and CalPred (blue) using simulated quantitative context with varying context-specific effects on PGS accuracy at confidence levels of **(Top)** 95% and **(Bottom)** 50%. Each simulation setting was repeated 30 times. We calculated the context-specific magnitude defined by ΔR^2^, the relative difference between R^2^ in the top and bottom decile, ranging from 0 to 150%. Alternating shading indicates different context-specific magnitude levels (ΔR^2^). Within each shaded block, the six boxplots (two methods across three strata) correspond to the same ΔR^2^ value.

### Context-specific calibration of PGS-based predictions in All of Us

Having used simulations to explore calibration of PGS-based prediction intervals, we next turned to real data using LDL cholesterol and BMI from the All of Us Research Program^14^ (AoU) as example traits. To minimize ancestry-driven context effects^5,15^, we restricted analyses to participants self-reporting as White (White SIRE; **Methods**) and focused on age, sex, reported income, and reported education as contextual factors. We observed a similar marginal (average) calibration in AoU to that reported by Xu et al. in the UK Biobank. Although both CalPred and PredInterval showed similarly calibrated 95% prediction intervals when averaged across all individuals (**Figure 3**), their calibration within specific contexts differed across methods. For LDL across age deciles, CalPred maintained 95% coverage, while PredInterval ranged from 95.0% to 97.8%, with similar results for BMI across sex, income, and education strata (**Figure 3**). These miscalibrations directly reflected PGS contextual accuracy **(Supplementary Figure 5)**: LDL PGS R^2^ varied from 0.29% (oldest decile) to 0.63% (youngest decile; overall R^2^ = 0.39%; ΔR^2^ = −87%). We replicated these findings using PGSs for LDL and BMI as previously trained in UK Biobank^6^ to confirm that CalPred remained well calibrated when using externally trained PGS **(Supplementary Figure 6)**.

**Figure 3.**
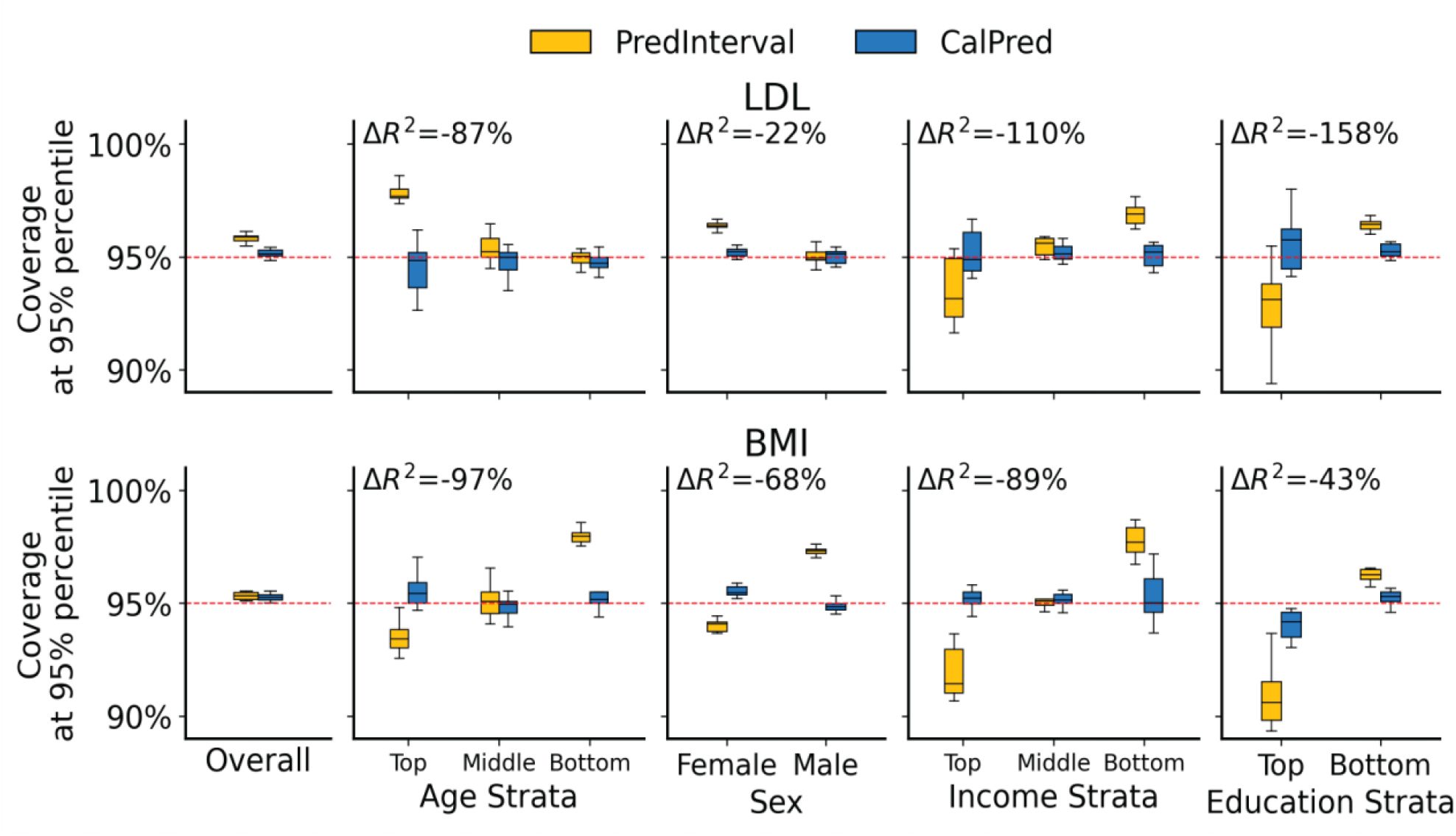
CalPred provides calibrated intervals for LDL and BMI across contexts in All of Us, whereas PredInterval failed. We compared the coverage between PredInterval (yellow) and CalPred (blue) on real data from the AoU^14^ White SIRE population for (Top) LDL and (Bottom) BMI trait. We reported the coverage at 95% prediction interval in the overall testing dataset and across age, sex, income, and education strata. For each trait-context pair, we annotated the context-specific effects ΔR^2^ calculated using PGSs trained within each CV and residualized phenotypes.

## Discussion

Taken together, our results show that marginal calibration is insufficient for PGS-based prediction intervals in real-world settings. CalPred achieves calibration across contexts, whereas PredInterval – despite guaranteeing marginal calibration through conformal prediction^16^ – is mis-calibrated within specific contexts strata when PGS performance exhibits contextual variability. We focused on contextual calibration within a single genetic ancestry to highlight the impact of non-ancestry contexts – we expect the miscalibration to be a larger problem when considering genetic ancestry impact on PGS performance. We also highlight that CalPred’s calibration data must be independent of training data to avoid overfitting.

Looking forward, we highlight the potential and utility of both CalPred and PredInterval as methods to estimate confidence intervals for PGS-based prediction: PredInterval as a non-parametric approach requiring minimal assumptions and CalPred as a parametric approach leveraging heteroskedastic regression to adjust for context to maintain calibration. Future works combining the two directions, e.g., by integrating conformal prediction into CalPred or reversely using heteroskedastic regressions to normalize data in PredInterval is a promising direction that could combine guaranteed marginal coverage with context-specific calibration. As genomic medicine increasingly serves diverse populations, context-specific calibration is needed for evaluating and deploying PGS-based prediction tools.

## Methods

### Simulations for no context-specific effect on PGS accuracy

We replicated the simulations described in Xu et al.^7^ (Figures 2,4; https://github.com/xuchang0201/PredInterval/blob/main/Manuscript/Simulations/Main_Simulations_Quantitative.R commit 276822c). All code and data necessary to reproduce the simulation results are available in Zenodo (see **Data Availability**). Genotypes were simulated for 200,000 individuals using HAPGEN2^17^ with linkage disequilibrium (LD) patterns from European populations in the 1,000 Genomes Project reference panel^18^. We restricted analyses to HapMap3^19^ variants on chromosome 1 (m = 82,204 SNPs). As described in Xu et al., we simulated h^2^ ∈ {0.2, 0.5, 0.8} and polygenicity ρ ∈ {0.001, 0.01, 0.1, 1}, where ρ denotes the proportion of causal variants and m_causal_ = ρm. Similarly, training datasets of size n_train_ ∈ {5000, 50000} and testing datasets of fixed size n_test_ = 10,000 were randomly sampled from the full cohort. Following Xu et al.’s simulation steps, we selected causal variants at random based on m_causal_, and their effect sizes were drawn independently from N(0, h^2^/m_causal_). Individuals’ genetic values (GV) were calculated as the dot product of genotypes and causal effect sizes. As described in Xu et al., we simulated phenotypes by adding noise drawn from N(0,1-h^2^) to the genetic values, then normalizing them to mean 0 and unit variance. Each simulation setting was repeated 30 times across 24 settings, yielding a total of 720 simulations.

For PredInterval, we followed the 5-fold cross-validation framework described in Xu et al^7^. In each fold, 4,000 or 40,000 samples were used to perform GWAS and construct polygenic scores (PGS), while the remaining 1,000 or 10,000 held-out samples were used to compute residuals and construct prediction intervals for the testing dataset. GWAS was performed using PLINK2^20^, and PGS was constructed using PRS-CS^21^.

For CalPred, we used the PGS trained on the first cross-validation fold of PredInterval and the corresponding held-out samples as an independent calibration dataset to fit the prediction model. This design ensured that CalPred had independent training and calibration datasets without requiring additional data, while maintaining identical training data provided for both methods for a fair comparison. CalPred*-overfit was implemented by fitting the prediction model on the same data used to train PGS, following the Xu et al. simulation code provided at: https://github.com/xuchang0201/PredInterval/blob/main/Manuscript/Benchmark_Comparison_Methods/CalPred.R commit d7350b0, line 39.

Following Xu et al, we evaluated the performance in identifying high-risk individuals across all 24 simulation settings. High-risk thresholds were defined as the 95th percentile of phenotypic values in the training dataset^7^. Ground-truth high-risk individuals in the testing dataset were those with phenotypic values greater than or equal to the threshold, and those with prediction upper bounds exceeding the threshold were classified as high-risk. The high-risk identification rate was defined as the proportion of ground-truth high-risk individuals correctly classified as high-risk by each method. We further introduced false-positive rates by quantifying the misclassification of low-risk individuals as high-risk. Low-risk thresholds were defined as the 5th, 25th, or 50th percentiles of phenotypic values in the training dataset. Ground-truth low-risk individuals were those with phenotypic values at or below the corresponding threshold, and the misclassification rate was defined as the proportion of such individuals whose prediction upper bounds exceeded the high-risk threshold.

### Simulations for context-specific effects on PGS accuracy

We extended the simulations above to introduce PGS performance that varies across contexts. Specifically, we simulated a quantitative context variable C∼N(0,1), independently for each individual. All code and data necessary to reproduce the simulation results are available in Zenodo (see **Data Availability**).

Individual-level genetic values (GV) were generated using the procedure described above. Briefly, we followed one simulation setting in Xu et al. by fixing h^2^ at 0.5 and polygenicity ρ at 1, with causal effect sizes drawn from the setting, and GV computed as the dot product of phenotypes and causal effect sizes. To introduce context-specific predictive performance while preserving marginal heritability, phenotypes were generated as Y ∼ N(GV, exp(β_0_ + γC)), where GV was normalized to mean 0 and unit variance. We had γ ∈ {0, 0.05, …, 0.95, 1}, which governs how strongly the residual variance scales with the context variable C. We set β_0_=log(1/h^2^-1) to achieve targeted marginal heritability in the absence of context effects (γ=0). Therefore, we simulated the scenario in which context affects the residual variance of phenotypes.

As in the previous section, following Xu et al., we randomly sampled 50,000 individuals for training and 10,000 for testing from the full cohort of 200,000. Simulation at each context-specific effect size γ was repeated 30 times. We used 5-fold cross-validation with PredInterval. In each fold, 40,000 samples were used to train PGS, while the held-out 10,000 samples were used to compute residuals and construct prediction intervals for the testing dataset. For CalPred, we used the PGS trained on the first cross-validation fold of PredInterval and the corresponding held-out samples as an independent calibration dataset to fit the CalPred prediction model, where PGS was used in the mean term, and C and C^2^ were included in the variance term. GWAS was performed using PLINK2^20^, and PGS was constructed using PRS-CS^21^. Following CalPred work, we quantified context-specific prediction accuracy of PGS using R^2^, the squared Spearman correlation between PGS and the residual phenotype regressed on the quantitative context C^6^. Simulated context-specific magnitude was summarized by ΔR^2^ = (R^2^_top_ - R^2^_bottom_)/R^2^, where R^2^_top_ and R^2^_bottom_ denote prediction R^2^ in the top and bottom deciles of the context variable C, respectively. We chose the range of ΔR^2^ values below 150% to reflect the empirical patterns identified in the CalPred^6^ work.

To evaluate whether incorporating contextual covariates affects PredInterval calibration, we extended the PredInterval framework to include context variables in the prediction model, following the description by Xu et al. in the discussion section. Specifically, within each cross-validation fold, we fit a linear model on the training data, including PGS, the quantitative context variable C, and C^2^, along with an intercept. We computed residuals on held-out samples as the absolute difference between observed phenotypes and their prediction mean from the fitted linear model. These residuals were aggregated across folds to construct the empirical residual distribution. For each test individual, we obtained prediction means by averaging the point estimates from the fitted linear models, including PGS, C, and C^2^, across five cross-validation folds. Following Xu et al., we constructed prediction intervals by adding and subtracting sampled residuals from the empirical distribution to the predicted mean. This implementation ensures that contextual covariates are incorporated into the prediction model as described in Xu et al.’s discussion section, consistent with the PredInterval framework, while preserving its nonparametric residual-based construction of prediction intervals.

### Real-data analysis in All of Us

We used the same genotype and phenotype data as in CalPred^6^ study. Briefly, we used All of Us^14^ (AoU) v7 microarray data after QC with PLINK2^20^ (--geno 0.05 --maf 0.001 --max-alleles 2 --rm-dup exclude-all --chr 1-22), retaining 1.2 million SNPs. Analyses were restricted to self-identified non-Hispanic White participants. We quantile-normalized the most recent measures of LDL (n=59,915) and BMI (n=129,525) for “White” self-identified race and ethnicity for downstream analyses. Summary-level results are available at the GitHub repository, see **Code Availability**.

Individuals were randomly permuted, and 10,000 individuals were held out as the test set; the remaining were used for training. PredInterval was implemented using the 5-fold cross-validation described in Xu et al^7^. Within each cross-validation fold, we performed GWAS by PLINK2^20^ on quantile-normalized phenotype with standardized covariates, including age, sex, and the top 10 genotype PCs. PGS were constructed using PRS-CS^21^. Residuals were computed from held-out samples in each fold and used to construct prediction intervals for evaluation. For CalPred, we used PGS weights trained on the first CV fold of PredInterval and used the held-out individuals from that fold as the calibration dataset. We quantified the context-specific effect of 11 contexts: age, sex, top two genotype PCs, BMI (when not the target trait), smoking, alcohol, employment, ‘education years’, income, and number of years living in the current address. We included the 11 contexts, along with age^2^, PC1^2^, and PC2^2^, as variance terms in the CalPred model. In the mean model, we included PGS, age, sex, age×sex, the top 10 genotype PCs, BMI (when not the target trait), smoking, alcohol use, employment, education years, income, years lived at current address, and interactions between PGS and each of the 11 context variables. For each trait, we repeated the experiment 10 times.

For completeness, we replicated our results using PGS weights trained in the UK Biobank^22^ to AoU individuals, as described in the CalPred^6^ paper. For PredInterval, within each cross-validation fold, externally derived PGS values were used directly to compute residuals without retraining in AoU. For CalPred, all individuals outside the test dataset were used for calibration. Other implementation details were consistent with the standard approach described in the previous paragraph. For each trait, we repeated the experiment 10 times. Context-specific prediction accuracy of PGS R^2^ in real-data analysis was defined as the squared Spearman correlation between PGS and the residual phenotype regressed on age, sex, age×sex, and the top 10 genotype PCs.

## Data Availability

Simulation data, scripts, pre-generated intermediate files, and full results are available at https://doi.org/10.5281/zenodo.18840380, which allow replication of all simulation analyses and Figures 1 and 2.

## Data Availability

Simulation data, scripts, pre-generated intermediate files, and full results are available at https://doi.org/10.5281/zenodo.18840380, which allow replication of all simulation analyses and Figures 1 and 2. All of Us individual-level genotypes and phenotypes are available through the application at https://www.researchallofus.org. 1000 Genomes Project high-coverage data could be downloaded from http://ftp.1000genomes.ebi.ac.uk/vol1/ftp/data_collections/1000G_2504_high_coverage/working/20220422_3202_phased_SNV_INDEL_SV.

## Code Availability

Scripts used for simulation and summary-level results for generating Figures 1-3 are available at https://github.com/zhuozshi/CalPred-PredInterval; to replicate all simulation analyses, see Zenodo in Data Availability. CalPred software v0.1.1 is available at https://github.com/KangchengHou/calpred, accessed in December 2025. PredInterval software (commit 6cadfb3) is available at https://github.com/xuchang0201/PredInterval, accessed in December 2025.

## Acknowledgements

We gratefully acknowledge All of Us participants for their contributions, without whom this research would not have been possible. We also thank the National Institutes of Health’s All of Us Research Program for making available the participant data examined in this study. This work was supported by the National Institute of Aging R01AG085518 and the National Institute of Mental Health R01MH115676. The content is solely the responsibility of the authors and does not necessarily represent the official views of the National Institutes of Health.

