## Supplementary Figures for "CalPred yields calibrated intervals for polygenic risk prediction"

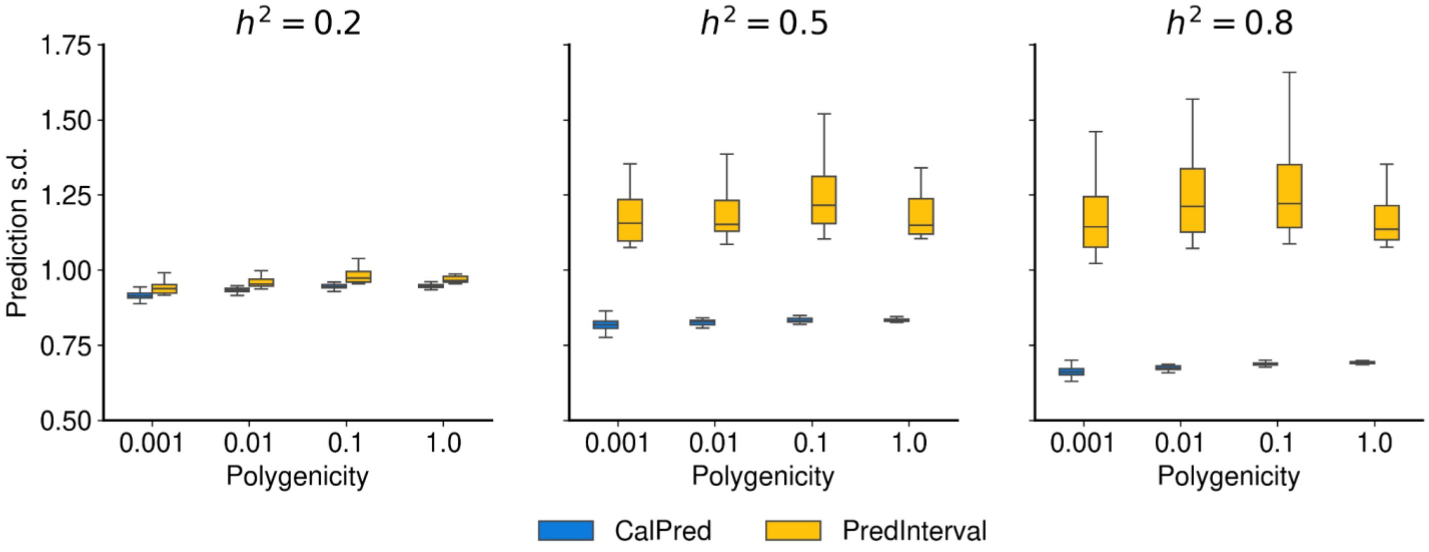


**Supplementary Figure 1. PredInterval produces wider prediction intervals compared to CalPred.** Comparison of prediction standard deviation (s.d.) between PredInterval (yellow) and CalPred (blue) across simulation settings with varying polygenicity = 0.001, 0.01, 0.1, and 1, and heritability = 0.2, 0.5, and 0.8.

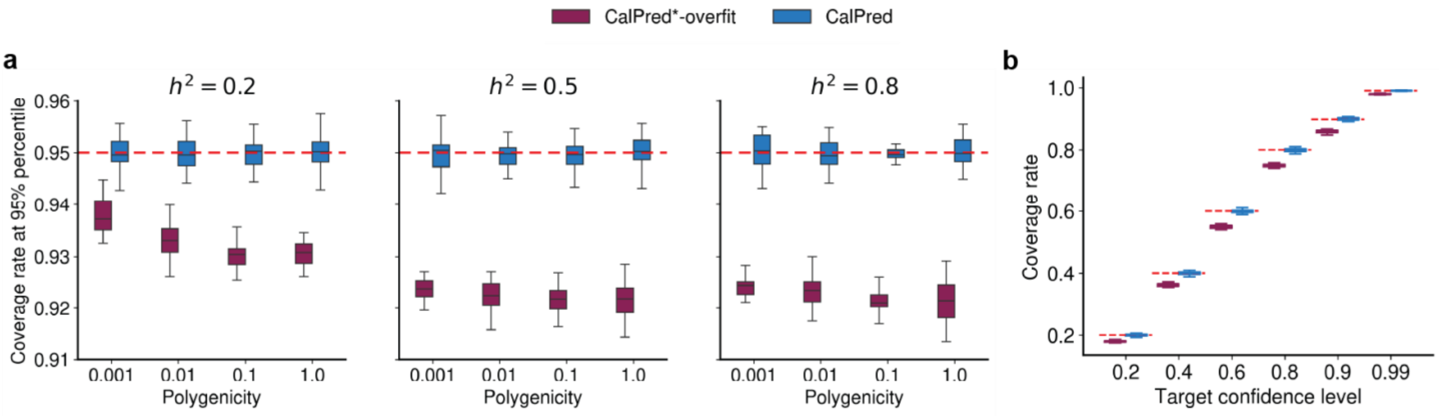


**Supplementary Figure 2. CalPred*(overfit) is mis-calibrated when training and calibration data is not separated.** Results are shown for simulated quantitative traits. Methods compared include CalPred (blue) and CalPred*(overfit) (purple), implemented with and without an independent calibration split from the training dataset, respectively. The test dataset size was 10,000 individuals, and each simulation setting was repeated 30 times. **a-b.** Results using a training dataset of size 50,000. The red dashed line indicates the targeted confidence level. **a.** Prediction coverage at the 95% confidence level across simulation settings with varying polygenicity = 0.001, 0.01, 0.1, and 1, and heritability = 0.2, 0.5, and 0.8. **b.** Coverage rates across targeted confidence levels 0.2, 0.4, 0.6, 0.8, 0.9, and 0.99, with heritability of 0.5 and polygenicity of 1.

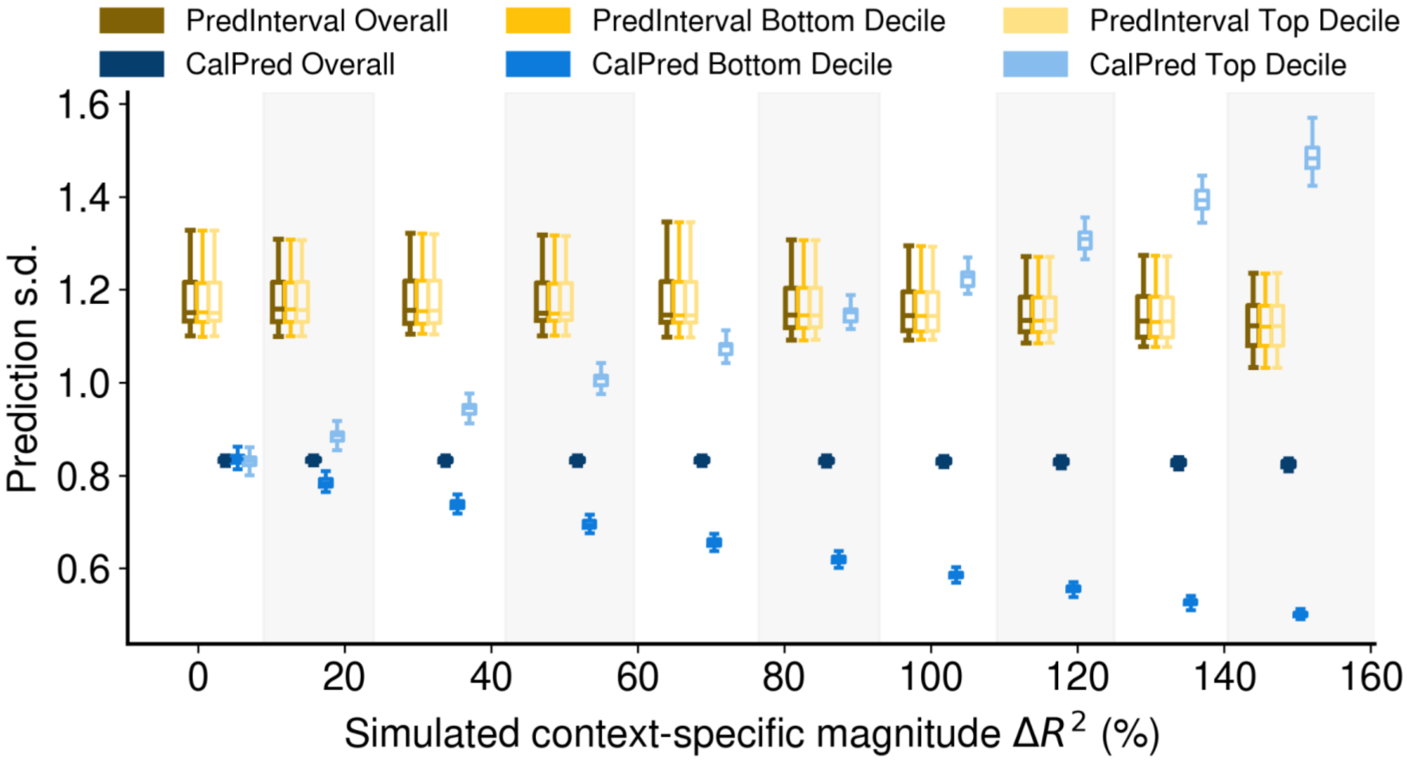


**Supplementary Figure 3. Miscalibration of PreInterval is driven by the static prediction standard deviation (s.d).** Comparison of prediction s.d. between PredInterval (yellow) and CalPred (blue) in the presence of a quantitative context and varying context-specific effects. Each simulation setting was repeated 30 times. Simulated context-specific magnitude is defined by $\Delta R^2$, the relative difference between $R^2$ in the top and bottom decile, ranging from 0 to 150%.

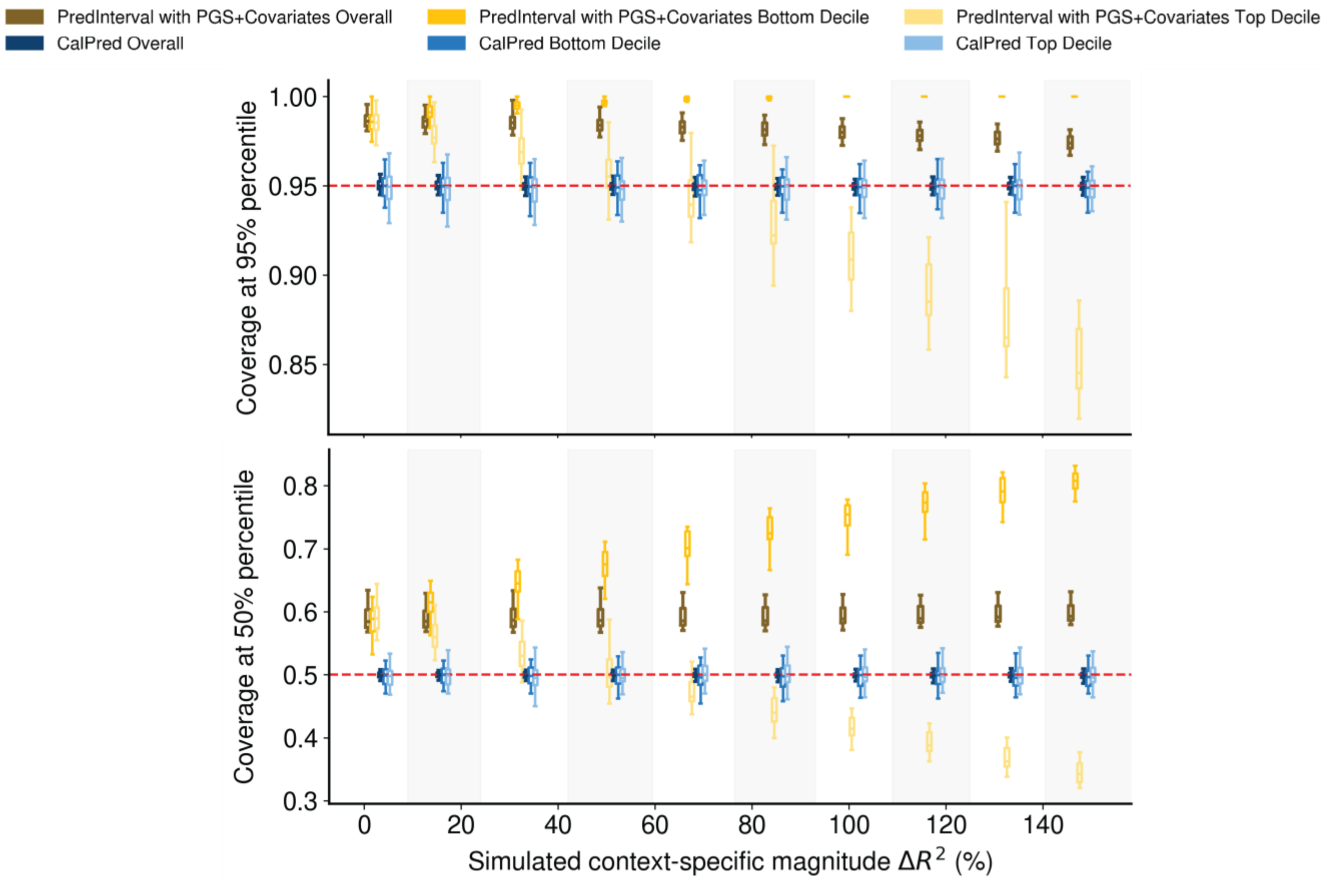


**Supplementary Figure 4. Miscalibration of PreInterval persists after including the context as a covariate.** We compared the coverages of PredInterval with PGS+covariates (yellow) and CalPred (blue) using simulated quantitative context with varying context-specific effects on PGS accuracy at confidence levels of **(Top)** 95% and **(Bottom)** 50%. Each simulation setting was repeated 30 times. We calculated the context-specific magnitude defined by $\Delta R^2$, the relative difference between $R^2$ in the top and bottom decile, ranging from 0 to 150%. Alternating shading indicates different context-specific magnitude levels ($\Delta R^2$). Within each shaded block, the six boxplots (two methods across three strata) correspond to the same $\Delta R^2$ value.

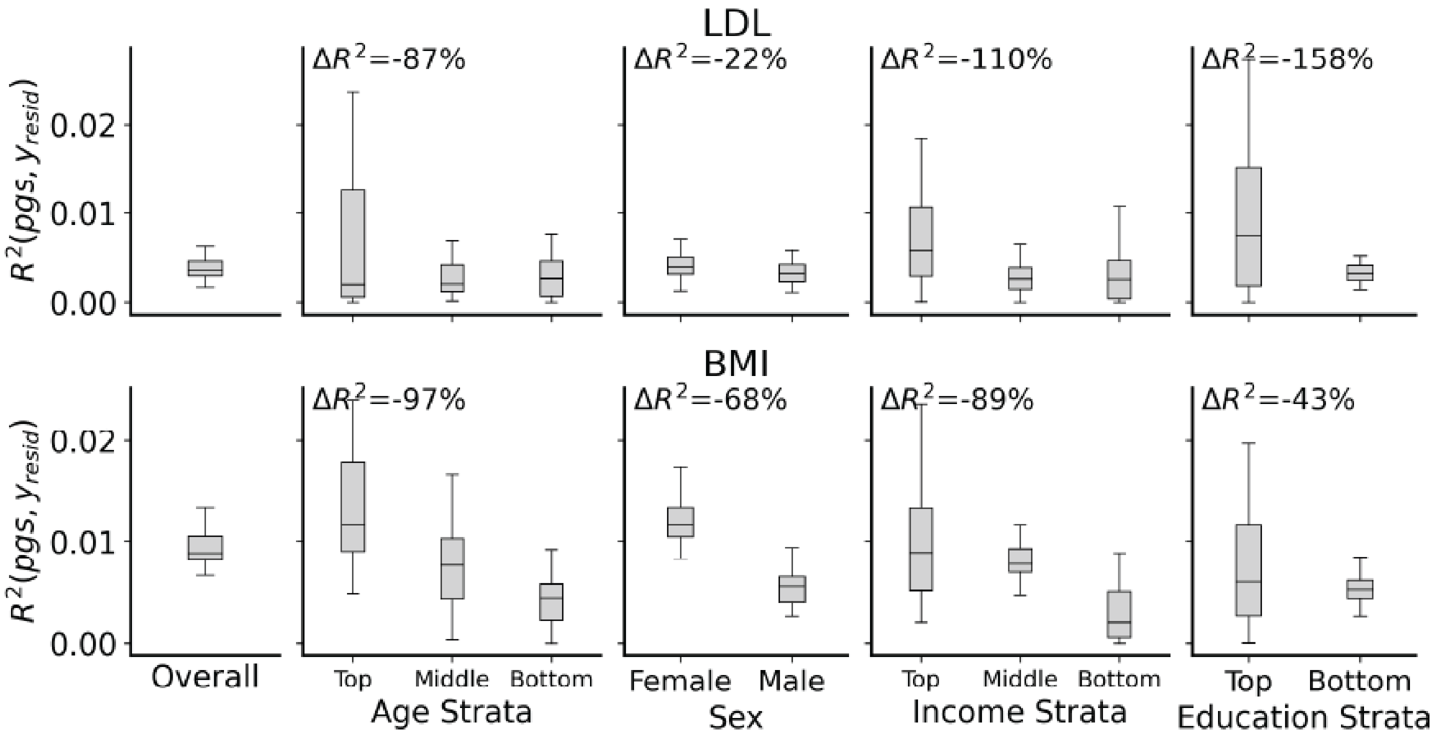


**Supplementary Figure 5. Context-specific prediction accuracy of PGS for LDL and BMI traits in All of Us. Top**: LDL. **Bottom**: BMI. Boxplots show the distribution of prediction $R^2$ between raw PGS and residualized phenotype evaluated within each subgroup defined by age, sex, income, and education strata. The relative change in prediction $R^2$ between the top and bottom subgroups for each context (context-specific magnitude $\Delta R^2$) is annotated at the top left of each panel.

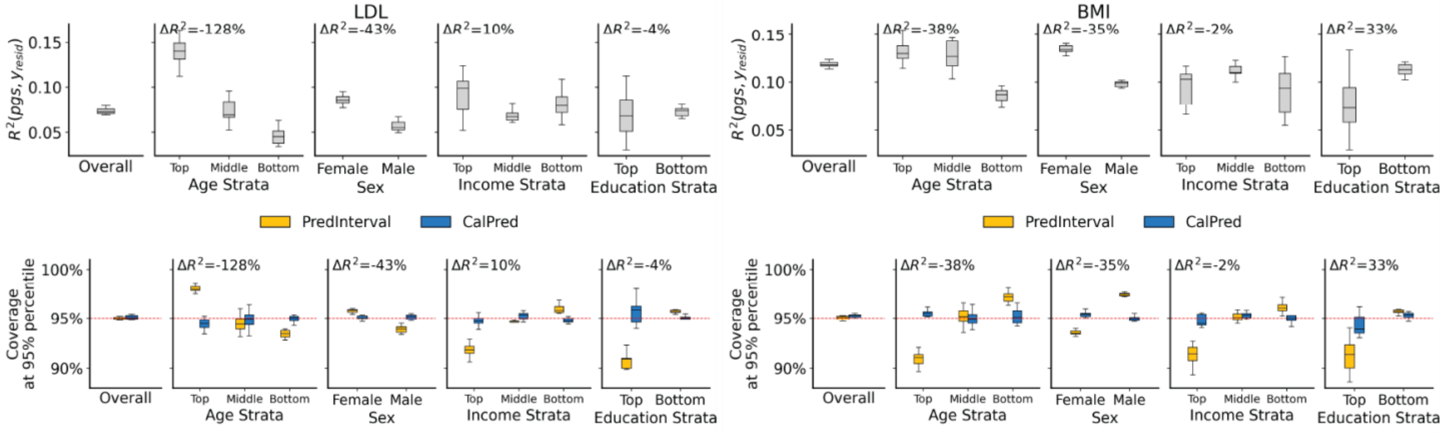


**Supplementary Figure 6. Context-specific prediction accuracy of PGS and calibration using PredInterval and CalPred for LDL and BMI traits in All of Us using PGS trained in UKBiobank.** (Top) Boxplots show the distribution of prediction $R^2$ between raw PGS and residualized phenotype evaluated within each subgroup defined by age, sex, income, and education strata. The relative change in prediction $R^2$ between the top and bottom subgroups for each context (context-specific magnitude $\Delta R^2$) is annotated at the top left of each panel. (Bottom)

Coverage of the prediction interval at 95% percentile. Context-specific prediction accuracy and coverage are illustrated in (**left**) LDL and (**right**) BMI in the All of Us v7 White SIRE population.
